# Changes in physical activity, sitting and sleep across the COVID-19 national lockdown period in Scotland

**DOI:** 10.1101/2020.11.05.20226381

**Authors:** Xanne Janssen, Leanne Fleming, Alison Kirk, Lesley Rollins, David Young, Madeleine Grealy, Bradley MacDonald, Paul Flowers, Lynn Williams

**Affiliations:** School of Psychological Sciences and Health, University of Strathclyde, Glasgow, UK; Department of Mathematics and Statistics, University of Strathclyde, Glasgow, UK

**Keywords:** SARS-COV-2, 24-hour movement behaviors, social distancing, covid-19, lockdown, behavior change

## Abstract

We examine the impact of the COVID-19 outbreak and concomitant restrictions (i.e. lockdown) on 24-hour movement behaviors (i.e. physical activity, sitting, sleep) in a purposive sample of people (n=3230) reporting change recruited on-line. Participants’ self-reported time spent in moderate-to-vigorous physical activity (MVPA), walking, sitting and sleep prior to lockdown (T1), during the first national lockdown (T2) and as restrictions initially started to ease (T3). For each 24-hour movement behavior, category-shifts are reported (positive, negative or did not change), as well as the percentage of participants recording positive/negative changes across clusters of behaviors and the percentage of participants recording improvement or maintenance of change across time. From T1 to T2 walking decreased, whereas MVPA, sitting and sleep increased, from T2 to T3 levels returned to pre-lockdown for all but MVPA. Participants who changed one behavior positively were more likely to report a positive change in another and 50% of those who reported positive changes from T1 to T2 maintained or improved further when restrictions started to ease. The current study showed that a large proportion of the sample reported positive changes, most notably those displaying initially poor levels of each behavior. These findings will inform salutogenic intervention development.

## 1. Introduction

The COVID-19 outbreak has affected people all around the world. In Scotland, a nationwide lockdown was announced on the 23rd of March [1]. This meant people were asked to stay at home and only go outside once a day for exercise, shopping for essential items, medical needs or to go to work if their job was vital. On the 28^th^ May the Scottish Government slowly started easing the lockdown measures meaning greater freedom, people could spent more time outside, meet up with friends and start to travel outside of their local area [2]. These rapid ongoing changes to people’s daily structure are likely to have influenced behavioural outcomes. However, whilst emerging research has explored the immediate impact of COVID-19, limited data exists to determine the lasting impact of these ongoing changes on behavioural outcomes.

Several studies have looked at the immediate effects of the COVID-19 pandemic on 24-hour movement behaviours (i.e. physical activity, sitting and sleep). Current evidence shows that behavioral patterns changed. Many people reported their physical activity declined, sedentary behavior increased and people reported poorer sleep [3,4]. Changes in 24-hour movement behaviours may further compound the negative changes reported in mental and physical health during the pandemic. Prior to the COVID-19 epidemic, it was already well documented that 24-hour movement behaviours are associated with physiological and mental health [5-7]. Since then, it has been suggested that changes in physical activity due to COVID-19 increase the risk of cardiovascular disease [8]. In addition, studies have reported that those who engaged in more physical activity, less sedentary behaviour and poorer sleep reported better mental health outcomes at the start of the covid-19 pandemic [4, 9].

While, evidence exists on changes in 24-hour movement behaviours at the start of the 1^st^ national lockdowns, very little is known about the maintenance of changes in these behaviours throughout the lockdown period and when restrictions initially started to ease. In addition, no study so far has examined the inter-relationship between these behaviours (e.g. if someone positively changes sleep, do they also increase physical activity or reduce sedentary behavior?). The inter-relationship between these behaviours is of particular importance as recent research has highlighted that how behaviours interact with each other influences health (e.g. the positive effects of appropriate levels of physical activity may diminish if sleep is poor)[10]. Understanding changes in these 24-hour movement behaviours during lockdown is crucial in attempt to support people to maintain or return to healthy lifestyles post COVID-19. Therefore, this study aims to examine changes in 24-hour movement behaviours from: 1) pre-lockdown (T1) to the 1^st^ UK national lockdown (T2) and 2) 1^st^ UK national lockdown to initial easing of lockdown (T3). The study aims to answer the key research questions:

1. How did the 24-hour movement behaviours change from T1 to T2 and from T2 to T3?
2. Was there evidence of changes in the 24-hour movement behaviours clustering together?
3. Were changes in the 24-hour movement behaviours maintained over time as lockdown restrictions eased?

## 2. Materials and Methods

### 2.1 Participants and procedure

Participants were primarily recruited through social media advertisements on Facebook and Twitter as part of the CATALYST study [11]. Participants were eligible to take part in the study if they were 18 years or older, currently residing in Scotland, and interested in sharing their experience of positive change. Interested participants were directed to an online survey on Qualtrics. Before completing the questionnaire, participants were asked to read the participant information sheet and sign the online consent form. Data collection took place in two phases. Phase 1 ran from 20th May 2020 to 12 June 2020 (spanning the 9th to the 12th week of national lockdown in Scotland), and phase 2 ran from 3rd August to 21st August 2020 (coinciding with continued easing of lockdown restrictions in Scotland). During phase 1, participants were asked to report on behavioral outcomes relating to life pre-lockdown (T1) and life during lockdown (T2). Phase 2 focused on behavioral outcomes as lockdown in Scotland was easing (T3). All materials and procedures were reviewed and approved by the University Ethics Committee and all participants gave informed consent.

### 2.2 Measures

#### 2.2.1 Behavioral outcomes

Physical activity was measured using six items from the short form version of the international physical activity questionnaire (IPAQ) which has acceptable measurement properties among diverse populations of 18-65y old adults [12]. Participants were asked to report the frequency and duration of moderate and vigorous physical activities as well as walking during a normal week pre-lockdown (T1), during the 1^st^ UK lockdown (T2) and as restrictions initially started to ease (T3). Time spent in moderate-to-vigorous physical activity (MVPA) and walking were calculated as minutes per week and for each behavior participants were classified in one of five groups: 1) engaging in less than 30 min of moderate-to-vigorous physical activity (MVPA)/ walking per week, 2) engaging in 30 to <60 minutes of MVPA/ walking per week, 3) engaging in 60 to <150 min of MVPA/ walking per week, 4) engaging in 150 to <300 min of MVPA/ walking per week, and 5) engaging in ≥300 min of MVPA/ walking per week [13]. Changes in MVPA and walking were categorized as positive if participants changed from a lower category to a higher category (e.g. from 30 min/week to 60 to <150 min/week), negative if the changed from a higher to a lower category (e.g. from 60 to <150min/week to <30 min/week), participants were categorized as no change if they stayed within the same category.

Sitting time (i.e. sedentary behavior) was measured using the last item from the short form version of the international physical activity questionnaire (IPAQ) [12]. Participants were asked to report the hours spent sitting during a normal week pre-lockdown (T1), during the 1^st^ UK lockdown (T2) and as restrictions initially started to ease (T3). Time spent sitting was classified into 4 groups: 1) seated <4 hours per day, 2) seated 4 to <6 hours per day, 3) seated 6 to <8 hours per day, and 4) seated ≥8 hours per day [14]. Changes in sitting were categorized as positive if participants changed from a higher category to a lower category (e.g. from ≥8 hr/day to 6 to <8 hours), negative if the changed from a higher to a lower category (e.g. from 6 to <8 hours to ≥8 hr/day), participants were categorized as no change if they stayed within the same category.

Sleep duration was measured by asking participants to report on the number of hours sleep they got at night pre-lockdown (T1), during the 1^st^ UK lockdown (T2) and as restrictions initially started to ease (T3). Participants were categorized into one of five categories at each time point, in line with the National Sleep Foundation ‘Sleep Duration Recommendations’ (Supplementary Table S1) [15]. These categories were: 1) insufficient sleep, 2) appropriate amount of sleep (low), 3) recommended amount of sleep, 4) appropriate amount of sleep (high) and 5) too much sleep. Positive changes in sleep across time points were categorized in the following ways:

- Change from ‘insufficient sleep’ or ‘too much sleep’ to ‘appropriate sleep’ (high or low)
- Change from ‘insufficient sleep’ or ‘too much sleep’ to ‘recommended sleep’
- Change from ‘appropriate sleep ‘(high or low) to ‘recommended sleep’

Negative changes in sleep across time points were categorized in the following ways:

- Change from ‘recommended sleep’ to any other category
- Change from appropriate (high/low) to ‘insufficient sleep’ or ‘too much sleep’

Participants were categorized as no change if they stayed within the same category or changed from ‘insufficient sleep’ to ‘too much sleep’ or ‘appropriate sleep (low)’ to ‘appropriate sleep (high)’ or vice versa.

### 2.3 Statistical analysis

Descriptive statistics for T1, T2 and T3 are provided for all outcomes. The first stage of this study focused on examining changes from T1 to T2, whereas in stage two we focused on changes from T2 to T3. Due to the two-stage nature of the study and to maximize sample size, paired t-tests were run to examine significant changes between T1 and T2, and T2 and T3. Reported p-values are therefore unadjusted for multiple comparisons. In addition, based on the categorical data described above, an overview of the number and percentage of participants recording a positive change, negative change, or no change in MVPA, walking, sitting and sleep from T1 to T2 and from T2 to T3 was provided. Maintenance of change was reported as the percentage of participants who reported a positive category change from T1 to T2 and maintained or further improved this change from T2 to T3. All analyses were done with SPSS (version 26) at a 5% significance level.

## 3. Results

Descriptive statistics for each time point are described in Table 1. In total 3230 participants took part in the study and reported on at least one outcome variable at T1. Participants were on average 46.2 years of age and the majority of the sample was female (80.4%).

**Table 1.** Descriptive statistics

|  | *Pre-lockdown (T1)* |  |  | *Lockdown (T2)* |  |  | *Easing of lockdown (T3)* |  |  |
| --- | --- | --- | --- | --- | --- | --- | --- | --- | --- |
|  | N | Mean | SD | N | Mean | SD | N | Mean | SD |
| Age | 3230 | 46.2 | 15.30 |  |  |  |  |  |  |
| Gender (% female) | 3209 | 80.5 |  |  |  |  |  |  |  |
| Sitting per day (min) | 2552 | 396.91 | 188.02 | 2340 | 427.42 | 210.86 | 1806 | 388.25 | 197.44 |
| MVPA per week (min) | 2526 | 368.82 | 535.50 | 2421 | 426.74 | 616.44 | 1822 | 404.63 | 506.38 |
| Walk per week (min) | 2695 | 478.60 | 521.43 | 2412 | 419.59 | 422.05 | 1841 | 487.45 | 488.87 |
| Sleep per night (hrs) | 3216 | 6.59 | 1.23 | 2473 | 6.88 | 1.55 | 1874 | 6.64 | 1.33 |

### 3.1 How did the 24-hour movement behaviors change from T1 to T2 and from T2 to T3?

Of the full sample 2458 participants completed T1 and T2 for at least one of the outcomes. Results of the paired-tests showed significant increases in sitting time (MD=29.10 min/day), MVPA (MD=68.40 min/week), sleep (MD=0.28 hr/day) and decreased their walking (MD=-55.58 min/week). Between T2 and T3, participants decreased their sitting time (MD)=-30.43 min/day), sleep (MD=-0.22 hr/day) and increased their walking (MD=60.89 min/week) (Table 2).

**Table 2.** Change in 24-hour movement behaviors over time

|  | N | Mean difference | 95% CI | p |
| --- | --- | --- | --- | --- |
| **T1 to T2** |  |  |  |  |
| *Sitting (min/day)* | 2297 | 29.1 | 21.96, 36.24 | <0.001 |
| *MVPA (min/week)* | 2358 | 68.40 | 45.73, 91.07 | <0.001 |
| *Walking (min/week)* | 2363 | -55.58 | -77.98, -33.18 | <0.001 |
| *Sleep (hrs/night)* | 2458 | 0.28 | 0.22, 0.34 | <0.001 |
| **T2 to T3** |  |  |  |  |
| *Sitting (min/day)* | 1495 | -30.43 | -39.27, -21.59 | <0.001 |
| *MVPA (min/week)* | 1548 | 0.13 | -29.06, 28.81 | 0.993 |
| *Walking (min/week)* | 1557 | 60.89 | 37.57, 84.21 | <0.001 |
| *Sleep (hrs/night)* | 1608 | -0.22 | -0.28, -0.15 | <0.001 |

When examining categorical shifts within each of the individual behaviors from T1 to T2 results showed 29.5% of the participants reported a positive change for MVPA, 25.4% were already in the highest category and maintained that position, 21.5% reported no change and 23.7% reported a negative change. For walking 23.9% of the participants reported a positive change, 38.0% were already in the highest category and maintained at that position, 10.6% reported no change and 27.5% reported a negative change. For sitting, 19.5% of the participants reported a positive change, 8.2% were already in the lowest category and maintained at that position, 44.8% reported no change and 27.8% reported a negative change. Last, for sleep 23.6% of the participants reported a positive change, 37.7% were already sleeping optimally and maintained that position, 17.9% reported no change and 20.8% reported a negative change (Supplementary Table S2).

### 3.2 Was there evidence of changes in the 24-hour movement behaviors clustering together?

Table 3 shows cross-behavioral changes from T1 to T2. Briefly, of those recording a positive change in physical activity (i.e. MVPA and walking) 37.88% reported a positive change in sleep and 17.17% reported a negative change, 34.85% reported a positive change in sitting and 14.14% reported a negative change. In total 16.67% reported a positive change in all four behaviors. Of those recording a negative change in physical activity, 21.35% reported a positive change in sleep and 31.77% reported a negative change, and 7.29% reported a positive change in sitting and 45.31% reported a negative change. In total 16.15% reported a negative change in all four behaviors.

**Table 3.**
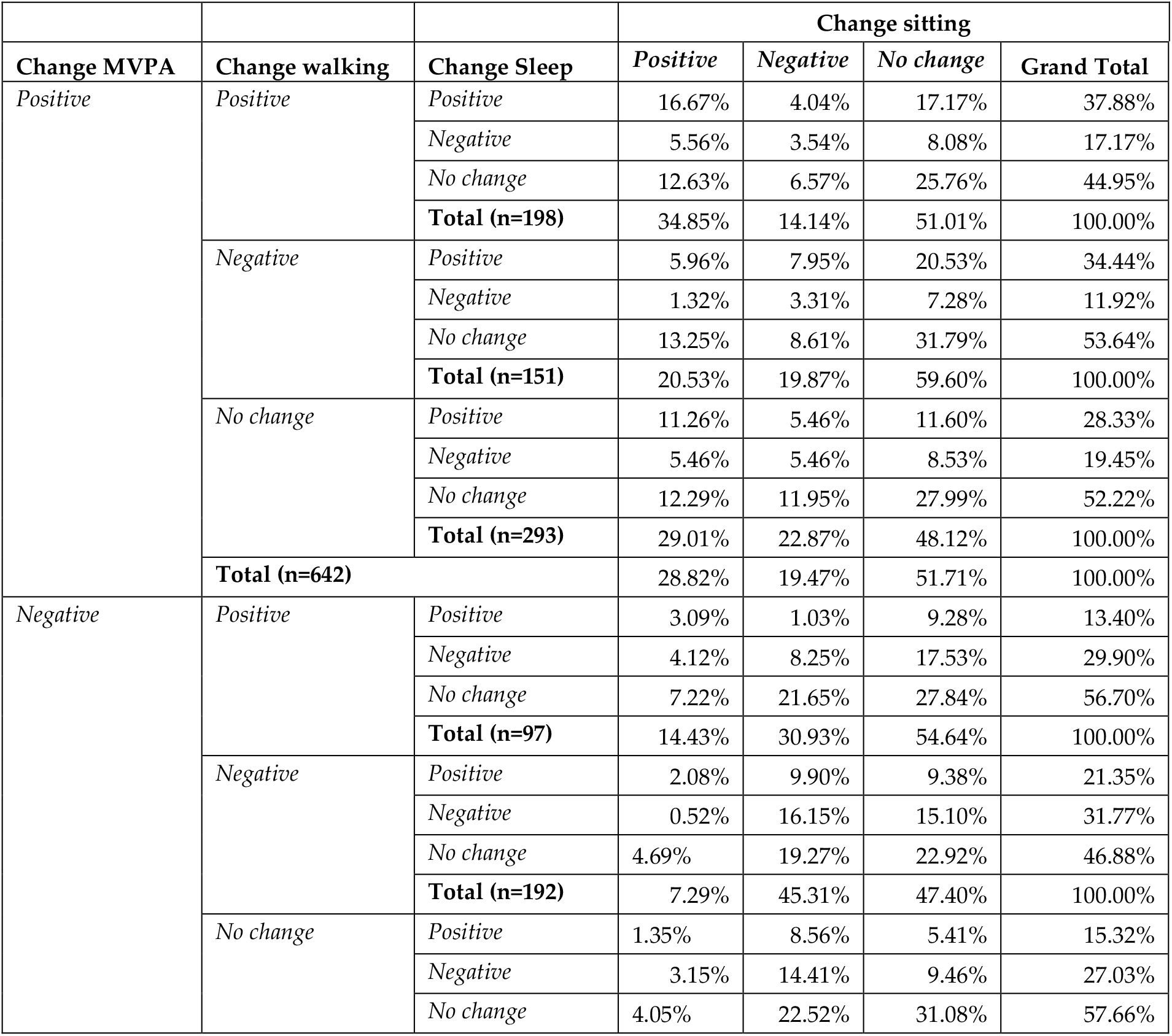
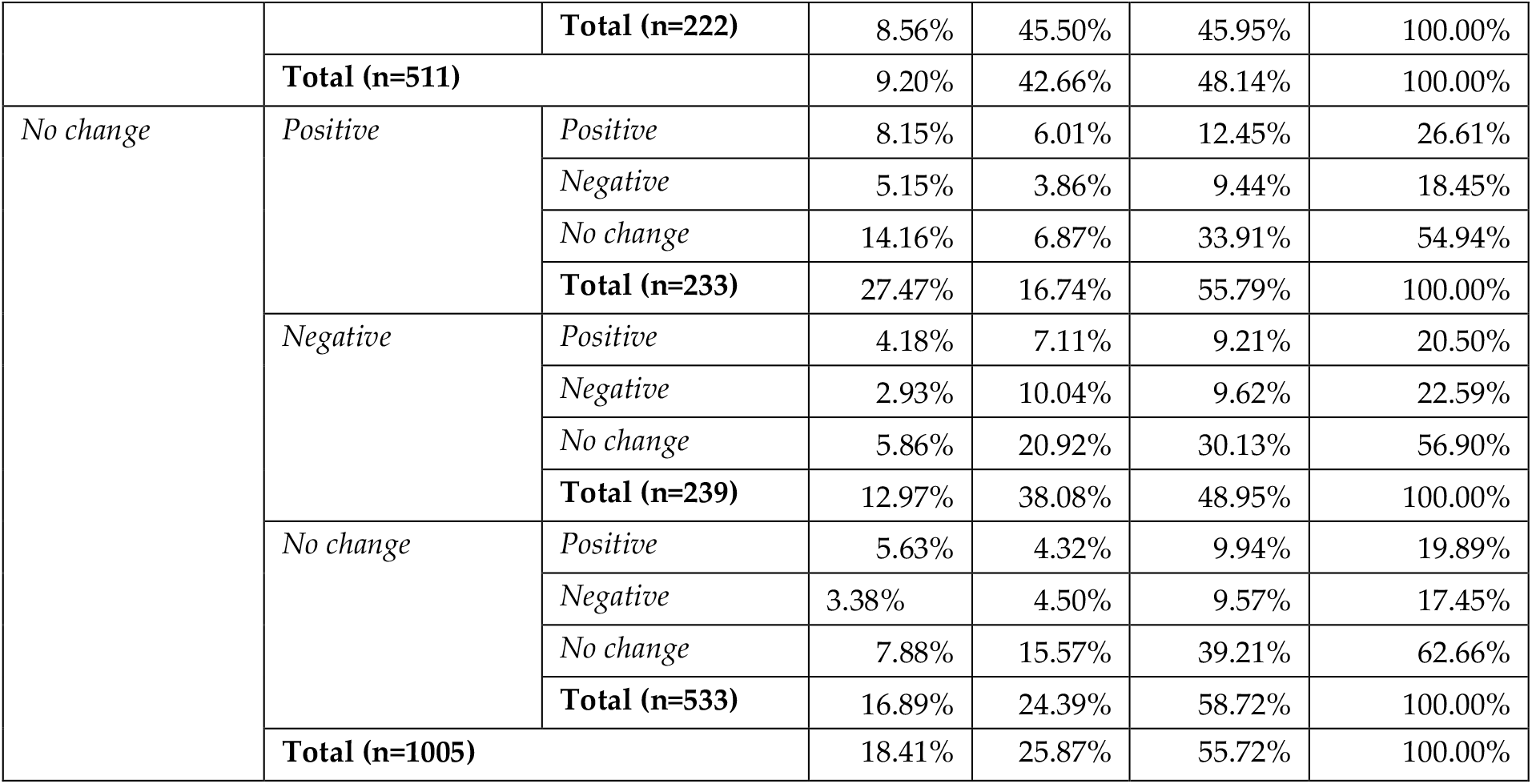
Cross-behavioral change from T1 to T2 (n=2158).

Table 4 shows cross-behavioral changes from T2 to T3. Briefly, of those recording a positive change in physical activity (i.e. MVPA and walking) 21.43% reported a positive change in sleep and 14.29% reported a negative change in sleep, 39.29% reported a positive change in sitting and 17.86% reported a negative change in sitting. In total, 10.71% reporting a positive change in all four behaviors. Of those recording a negative change in physical activity, 26.09% reported a positive change in sleep and 17.39% reported a negative change in sleep and 8.70% reported a positive change in sitting and 26.09% reported a negative change. One participant (2.17%) reported a negative change in all four behaviors.

**Table 4.** Cross-behavioral change from T2 to T3 (n=1413).

| **Change MVPA** | **Change walking** | **Change Sleep** | **Change sitting** |  |  |  |
| --- | --- | --- | --- | --- | --- | --- |
|  |  |  | *Positive* | *Negative* | *No change* | **Grand Total** |
| *Positive* | *Positive* | *Positive* | 10.71% | 3.57% | 7.14% | 21.43% |
|  |  | *Negative* | 7.14% | 1.79% | 5.36% | 14.29% |
|  |  | *No change* | 21.43% | 12.50% | 30.36% | 64.29% |
|  |  | **Total (n=56)** | 39.29% | 17.86% | 42.86% | 100.00% |
|  | *Negative* | *Positive* |  |  | 15.38% | 15.38% |
|  |  | *Negative* | 7.69% |  | 17.95% | 25.64% |
|  |  | *No change* | 20.51% | 7.69% | 30.77% | 58.97% |
|  |  | **Total (n=39)** | 28.21% | 7.69% | 64.10% | 100.00% |
|  | *No change* | *Positive* | 6.75% | 1.98% | 8.33% | 17.06% |
|  |  | *Negative* | 8.33% | 3.17% | 7.14% | 18.65% |
|  |  | *No change* | 25.00% | 9.92% | 29.37% | 64.29% |
|  |  | **Total (n=252)** | 40.08% | 15.08% | 44.84% | 100.00% |
|  | **Total (n=347)** |  | 38.62% | 14.70% | 46.69% | 100.00% |
| *Negative* | *Positive* | *Positive* | 4.00% |  | 8.00% | 12.00% |
|  |  | Negative | 2.00% | 4.00% | 18.00% | 24.00% |
|  |  | No change | 12.00% | 14.00% | 38.00% | 64.00% |
|  |  | **Total (n=50)** | 18.00% | 18.00% | 64.00% | 100.00% |
|  | Negative | Positive | 2.17% | 6.52% | 17.39% | 26.09% |
|  |  | Negative | 2.17% | 4.35% | 10.87% | 17.39% |
|  |  | No change | 4.35% | 15.22% | 36.96% | 56.52% |
|  |  | **Total (n=46)** | 8.70% | 26.09% | 65.22% | 100.00% |
|  | No change | Positive | 3.98% | 4.42% | 9.73% | 18.14% |
|  |  | Negative | 5.31% | 5.31% | 12.83% | 23.45% |
|  |  | No change | 15.49% | 15.04% | 27.88% | 58.41% |
|  |  | **Total (n=226)** | 24.78% | 24.78% | 50.44% | 100.00% |
|  | **Total (n=322)** |  | 21.43% | 23.91% | 54.66% | 100.00% |
| No change | Positive132 | Positive | 3.79% | 3.03% | 6.82% | 13.64% |
|  |  | Negative | 5.30% | 3.79% | 9.85% | 18.94% |
|  |  | No change | 15.15% | 11.36% | 40.91% | 67.42% |
|  |  | **Total (n=132)** | 24.24% | 18.18% | 57.58% | 100.00% |
|  | Negative | Positive | 8.06% | 3.23% | 14.52% | 25.81% |
|  |  | Negative | 6.45% |  | 3.23% | 9.68% |
|  |  | No change | 17.74% | 14.52% | 32.26% | 64.52% |
|  |  | **Total (n=62)** | 32.26% | 17.74% | 50.00% | 100.00% |
|  | No change | Positive | 4.55% | 3.27% | 8.36% | 16.18% |
|  |  | Negative | 5.27% | 4.00% | 7.27% | 16.55% |
|  |  | No change | 20.00% | 12.91% | 34.36% | 67.27% |
|  |  | **Total (n=550)** | 29.82% | 20.18% | 50.00% | 100.00% |
|  | **Total (n=744)** |  | 29.03% | 19.62% | 51.34% | 100.00% |

### 3.3 Were changes in the 24-hour movement behaviors maintained over time as lockdown restrictions eased?

When examining categorical shifts within each of the individual behaviors from T2 to T3 results showed 25.1% of the participants reported a positive change for MVPA, 31.3% were already in the highest category and maintained at that position, 21.3% reported no change and 22.3% reported a negative change. For walking 23.7% of the participants reported a positive change, 42.5% were already in the highest category and maintained at that position, 11.4% reported no change and 22.5% reported a negative change. For sitting 29.8% of the participants reported a positive change, 8.4% were already in the lowest category and maintained at that position, 42.6% reported no change and 19.2% reported a negative change. Last, for sleep 17.0% of the participants reported a positive change, 43.7% were already in the optimal sleep category and maintained at that position, 21.1% reported no change and 18.2% reported a negative change (Supplementary Table S3).

Of those reporting data at T1, T2 and T3, 436 participants reported a positive category change in MVPA from T1 to T2, 372 participants reported a positive category change in walking, 440 participants reported a positive category change in sitting and 370 a positive category change in sleep. Of these participants 57.6%, 63.2%, 49.8% and 56.5% maintained or improved their MVPA, walking, sitting and sleep from T2 to T3.

## 4. Discussion

The present study is the first to report on changes in 24-hour movement behaviors both during lockdown and when restrictions initially started to ease. Results showed participants decreased walking and increased MVPA, sleep and sitting from pre-lockdown to lockdown. However, when restrictions eased, walking, sleep and sitting returned to pre-lockdown levels. In addition, when examining categorical shifts within each of the individual behaviors, the percentage of participants recording a positive or negative change from T1 to T2 was very similar (20-30% of the sample for each behavior). The highest proportion of positive changers was found in those who, at T1, participated between 30-60 minutes of MVPA or walking per week (62.2% and 77.9%, respectively), 6 to <8h of sitting (29.2%) and either recorded insufficient or too much sleep (53.5% and 50%, respectively). Similar patterns were found when restrictions eased.

It has been reported that the benefit of a positive change in physical activity and sitting is highest in those initially reporting lower levels of physical activity and higher levels of sitting [16]. Interestingly, as mentioned, our study found a greater percentage of those initially displaying poor levels of each behavior reporting positive changes compared to those close to or meeting physical activity and sleep guidelines. This shows the “lockdown” intervention has great potential for targeting change in those that need it most. While, understandably, a lockdown approach to improve lifestyle behaviors is unwarranted, future studies should aim to gain a better understanding about what enabled those displaying poor levels of each behavior to change positively during lockdown through qualitative studies. Understanding the barriers and facilitators of positive change during lockdown enables researchers, practitioners and policy makers to provide opportunities for healthy behavior change for those most at risk in the post-COVID era.

Previous studies focusing on 24-hour movement behaviors during the COVID-19 pandemic have mainly reported immediate changes in these behaviors during the initial national lockdowns [3-4]. The results of the current study align with previously reported changes in physical activity, sitting and sleep. However, the current study also showed there is value in examining changes at a more in depth level. While, summary results show either an increase or decrease in these behaviors, when looking at percentages of change we found that the number of people increasing or decreasing a behavior was equal. Similarly, while summary results show levels of physical activity, sitting and sleep returned to pre-COVID levels, 50% of the positive changers maintained their change. Understanding what enabled these people to positively change and then maintain their positive health behaviors is crucial in developing interventions and policies which enables others to do the same.

The results in the current study also shows the interactive nature of the relationship between the three different movement behaviors (i.e. physical activity, sitting and sleep). If one behavior changed positively participants were more likely to also record a positive change in at least one other behavior from pre-lockdown to lockdown. This is in line with previous research which has identified clustering of health behaviors [17]. In addition, this finding is especially important, as it has been shown that for many lifestyle behaviors the accumulative influence on health is larger than the sum of the two parts [18]. This may indicate that if someone shows a positive change in physical activity and sleep, the combined health benefit of these changes may be larger.

As mentioned one of the main strengths of this study is its longitudinal nature and large sample size. However, the current study was not without its limitations. First, participants were asked to self-report levels of physical activity, sitting and sleep, this may have led to self-report bias. Second, the sample was predominantly female, and some important socio-demographic groups are under-represented within the sample (e.g. BAME groups) [10], and therefore results cannot be generalized to the larger Scottish population.

## 5. Conclusions

The current study reports evidence on changes in physical activity, sitting and sleep over a prolonged period during the COVID-19 epidemic. Results showed that a large proportion of the sample reported positive changes, most notably those initially engaging in poor levels of each behavior. Future studies should examine the determinants of these positive changes to inform intervention development. Last, this study showed the inter-relationship between behaviors, therefore, it may be particularly important to examine the combined effects of healthy 24-hour movement behaviors in adults and develop strategies to target these behaviors collectively in order to maximize health benefit.

## Supplementary Materials

The following are available online at www.mdpi.com/xxx/s1, Table S1: Sleep recommendations categorization; Table S2. Number of participants in each movement behaviour category at T1 (rows) and T2 (columns); Table S3. Number of participants in each movement behavior category at T2 (rows) and T3 (columns).

## Data Availability

The data will be available on publication of the paper on the Open Science Framework http://doi.org/10.17605/OSF.IO/NWH48

## Supplementary Tables

**Table S1.** Sleep recommendations categorization

|  | *Insufficient* | *Appropriate*<br>*(low)* | *Recommended* | *Appropriate*<br>*(high)* | *Too much* |
| --- | --- | --- | --- | --- | --- |
| *18-25 year olds* | < 6 hours | 6 hours | 7 to 9 hours | 10 to 11 hours | ≥ 12 hours |
| *26-64 year olds* | < 6 hours | 6 hours | 7 to 9 hours | 10 hours | ≥ 11 |
| *+65 year olds* | < 5 hours | 5 to 6 hours | 7 to 8 hours | 9 hours | ≥ 10 |

**Table S2.** Number of participants in each movement behavior category at T1 (rows) and T2 (columns).

| **MVPA (min/week)** |  |  |  |  |  |  |  |
| --- | --- | --- | --- | --- | --- | --- | --- |
|  | <30 | 30 to <60 | 60 to <150 | 150 to <300 | 300+ | Total | Positive changers (%) |
| <30 | 265 | 14 | 72 | 61 | 129 | 541 | 51.02 |
| 30 to <60 | 9 | 8 | 11 | 12 | 5 | 45 | 62.22 |
| 60 to <150 | 81 | 10 | 106 | 82 | 117 | 396 | 49.74 |
| 150 to <300 | 64 | 7 | 60 | 127 | 193 | 451 | 42.79 |
| +300 | 94 | 16 | 95 | 120 | 600 | 925 | NA |
| Total | 513 | 55 | 344 | 402 | 1044 | 2358 |  |
| **Walk (min/week)** |  |  |  |  |  |  |  |
|  | <30 | 30 to <60 | 60 to <150 | 150 to <300 | 300+ | Total | Positive changers (%) |
| <30 | 45 | 7 | 27 | 23 | 52 | 154 | 70.78 |
| 30 to <60 | 10 | 5 | 21 | 16 | 16 | 68 | 77.94 |
| 60 to <150 | 36 | 9 | 78 | 60 | 156 | 339 | 63.72 |
| 150 to <300 | 28 | 16 | 79 | 122 | 187 | 432 | 43.3 |
| 300+ | 106 | 27 | 162 | 177 | 898 | 1370 | NA |
| Total | 225 | 64 | 367 | 398 | 1309 | 2363 |  |
| **Sitting (hr/day)** |  |  |  |  |  |  |  |
|  | <4 | 4 to <6 | 6 to <8 | 8+ |  | Total | Positive changers (%) |
| <4 | 188 | 120 | 61 | 36 |  | 405 | NA |
| 4 to <6 | 86 | 232 | 118 | 120 |  | 556 | 15.47 |
| 6 to <8 | 46 | 96 | 160 | 184 |  | 486 | 29.22 |
| 8+ | 35 | 76 | 101 | 638 |  | 850 | 24.94 |
| Total | 355 | 524 | 440 | 978 |  | 2297 |  |
| **Sleep (hr/night)** |  |  |  |  |  |  |  |
|  | *Insufficient* | *Appropriate (low)* | *Recommended* | *Appropriate (high)* | *Too much* | Total | Positive changers (%) |
| *Insufficient* | 157 | 59 | 122 | 3 | 3 | 344 | 53.49 |
| *Appropriate (low)* | 128 | 251 | 385 | 17 | 4 | 785 | 49.04 |
| *Recommended* | 122 | 199 | 924 | 48 | 6 | 1299 | NA |
| *Appropriate (high)* | 0 | 1 | 6 | 9 | 3 | 19 | 31.58 |
| *Too much* | 0 | 0 | 2 | 1 | 1 | 4 | 50.00 |
| Total | 407 | 509 | 1439 | 78 | 17 | 2451 |  |

Footnote: a positive change is illustrated in green, a negative change is illustrated in orange and no change is illustrated in yellow.

**Table S3.**
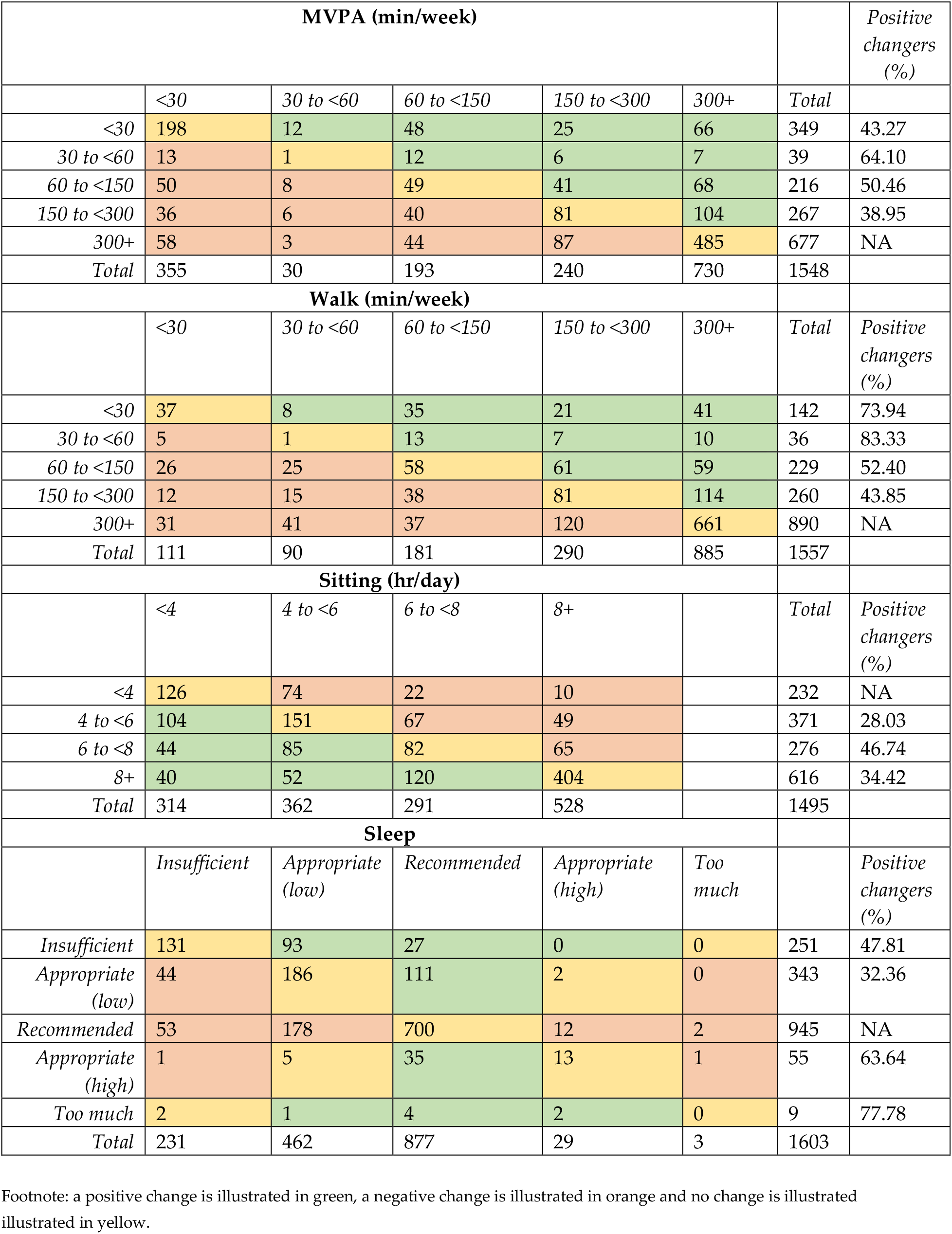
Number of participants in each movement behavior category at T2 (rows) and T3 (columns).

